# Implementing rapid pan-microbial metagenomics in paediatric intensive care

**DOI:** 10.1101/2025.10.07.25337257

**Authors:** Robbie Hammond, Angelika Kopec, Adela Alcolea-Medina, Rahul Batra, Katherine Brown, Virve Enne, Jonathan Edgeworth, Joanne Hemingway, Aparna Hoskote, Oscar Enrique Torres Montaguth, Sofia Morfopoulou, Mark Peters, Hamish Robertson, Samiran Ray, Anna Schmidt, Luke B. Snell, Nathaniel Storey, Daniel Ward, Surjo De, Garth Dixon, James Hatcher, James Soothill, Judith Breuer, Julianne R Brown

**Affiliations:** Microbiology, Virology and Infection Control, Great Ormond Street Hospital for Children, NHS Foundation Trust, London, UK; Intensive Care, Great Ormond Street Hospital for Children, NHS Foundation Trust, London, UK; Department of Infection, Immunity and Inflammation, University College London (UCL) Great Ormond Street Institute of Child Health, London, UK; Centre for Clinical Infection & Diagnostics Research, Guy’s & St. Thomas’ NHS Foundation Trust, London, UK

**Keywords:** clinical metagenomics, paediatrics, respiratory infection, intensive care, metagenomic sequencing, antimicrobial stewardship

## Abstract

**Background:** Clinical metagenomics studies have rarely reported rapid results with clinical impact, or deployment of protocols to centres beyond where they were developed. We describe the implementation of a previously validated rapid pan-microbial respiratory metagenomics service to paediatric intensive care, reporting modifications, feasibility, performance, clinical impact, and staff views.

**Methods:** Single-centre prospective interventional study of a metagenomics service testing respiratory samples from paediatric ICU patients at a London hospital between November 2024 and February 2025. This was based on a well-established published workflow detecting bacteria, fungi, and DNA/RNA viruses, with same-day preliminary results and next-day final results. Performance was assessed against routine microbiology. Clinical impact (change in antimicrobials, diagnosis or infection control) was evaluated through electronic health record review. Clinical staff were surveyed via an anonymous online semi-structured questionnaire.

**Findings:** 177 samples from 122 patients were tested. 174/177 (98%) passed quality control. Sensitivity at 16 hours sequencing was 89% (95% CI 80-95) for bacteria, 100% (95% CI 54-100) for fungi, and 87% (95% CI 76-95) for viruses, with specificities ≥99% across all kingdoms. Metagenomics identified 50 additional pathogens in 42/174 (24%) samples, all confirmed by orthogonal PCR testing. Clinical impact occurred in 51/174 (29.3%) samples, consisting of antimicrobial changes in 46/174 (26.4%), including 16/174 (9.2%) cessations and 10/174 (5.7%) initiations, and four samples (2.3%) with infection control implications. Among ICU staff surveyed (n=33), 72% viewed metagenomics positively, 0% viewing it negatively, and 67% perceived additional benefit over routine testing.

**Interpretation:** This clinical pilot of rapid respiratory metagenomics in paediatric intensive care demonstrated high specificity and good sensitivity. Protocols developed for an adult population were adapted to a paediatric population, including for polymicrobial upper respiratory tract samples that are more common for children. However, increased sensitivity and AMR prediction will be required to be truly disruptive. Whilst changes in patient management were observed, particularly for antimicrobial stewardship, delineating the true contribution of mNGS results is difficult in this study. Multi-centre studies over multiple winter seasons, with control arms and rationalising of patient cohorts and samples, are needed to assess impact on patient outcomes and cost-effectiveness.

**Research in Context:** *Evidence before this study:* Metagenomic next generation sequencing (mNGS) is increasingly being used in clinical settings for the diagnosis of infection. Technical performance of mNGS versus routine testing is frequently reported, but few studies report clinical impact of mNGS. Even rarer are studies reporting the adaptation of an established protocol from the centre of its development to a new centre with a different patient population (paediatric). We searched PubMed for studies published between 1st Jan 2020 and 1st Jan 2025, using the terms “paediatrics” AND “metagenomics” AND “treatment” AND either “respiratory” OR “pneumonia”. This search returned 135 abstracts, of which three were prospective studies using respiratory metagenomic testing in a fashion which impacted patient care. Of these, one study was not configured to detect RNA viruses. The remaining two studies reported 318 paediatric cases undergoing metagenomics. Neither study reported same day reporting of results with the intention of providing actionable results in real time. In summary there is minimal literature on the use of metagenomics as a rapid diagnostic test for respiratory infections in paediatric intensive care, or of adapting adult metagenomic protocols to paediatric populations.

*Added value of this study:* This observational prospective study is the first to show adaptation of a same day mNGS protocol able to detect bacteria, fungi, DNA and RNA viruses in a paediatric population. We show that modification of the adult protocol to include more upper respiratory and polymicrobial specimens was successful in achieving comparable performance standards. Metagenomics more rapidly identified bacterial pathogens which were also detected by standard-of-care methods as well as additional organisms that were not identified by routine methods. Taken together the data shows impact on antimicrobial prescribing, which could have implications for antimicrobial stewardship and clinical outcomes, although currently the precise contribution of mNGS to these antimicrobial prescribing decisions is unclear.

*Implications of all the available evidence:* This data suggests a role for respiratory metagenomics for diagnosing respiratory infection in paediatric intensive care. The results show the feasibility of implementing adult protocols in paediatric populations. Randomised control trials and cost-benefit analyses are still needed to assess true impact on length of hospital stay, clinical outcomes, antimicrobial usage and mortality.

## Introduction

Lower respiratory tract infections (LRTIs) cause a huge burden of paediatric disease, estimated at 502,000 deaths worldwide in children under 5 years.^1^ Aetiology can vary across a broad range of pathogens including bacteria, viruses, and fungi, leading to difficulty in diagnosis especially in critically unwell patients where infections are often polymicrobial.

Current routine LRTI diagnostics are usually a combination of culture based methods and multiplex PCR, with limitations around speed, sensitivity, and breadth of organisms tested.^2,3^ Metagenomics, the sequencing of all genetic material in a clinical sample, offers the potential of improving diagnostic yield due to its ability to potentially identify any organism in a sample.^4,5^ Furthermore, the pathogen-agnostic nature of metagenomics lends itself to diagnosing rare, unexpected or even novel organisms, making it an ideal test for pandemic preparedness and identifying novel infectious causes of syndromes. The utility of this has already been seen in children in helping identify the link between the increase in unexplained acute hepatitis in children and Adeno-associated virus 2, and in central nervous system infections.^6–10^

There has been recent interest in the potential of respiratory metagenomics as a first-line test for pneumonia in intensive care units,^3^ 6-hour wet-lab workflow using nanopore technology.^6^ This has then been piloted as a same-day service in Intensive Care Unit (ICU) patients in a single adult centre,^7,8^ with promising results and evidence of impact on patient care.

A question leading from these results will be the ability to adapt this workflow to different centres and populations, especially in paediatrics given the differences in respiratory microbiome, respiratory infection burden and immune function. Recent observational and surveillance studies have demonstrated potential utility for respiratory metagenomics in acute care ICU populations,^9–11^ but have not evaluated specifically in children nor a same-day clinical service.

In this paper, we assess feasibility, technical performance and clinical utility when implementing this modified protocol (previously only evaluated in parent centre with an adult population), as a same-day daily clinical service in a single paediatric centre.

## Methods

### Location & Sampling

The new service was run as standard of care on two paediatric ICU wards at a tertiary referral paediatric hospital; a general paediatric ICU and a cardiac paediatric ICU including an extracorporeal membrane oxygenation service (ECMO). The service began on 4^th^ November 2024, and ran Monday to Friday, with the data reported here having been collected for 17 weeks, with a two-week break from 23^rd^ December to 6^th^ January.

Patient samples were sent for all emergency admissions, or from patients selected by intensive care staff based on clinical assessment of patients with suspicion of LRTI. All respiratory sample types including broncho-alveolar lavage, endotracheal secretions, sputum, nasopharyngeal aspirate and pleural fluid were accepted. Samples had to be received by 9am to be run and return preliminary results on the same day, although if clinical staff communicated a delay in sample collection, runs could be delayed until 10.30am.

Separate aliquots of specimen were sent for metagenomics, culture and PCR; all three were not always sent as the decision was at the discretion of the clinician managing a patient. PCR testing was performed using a rapid commercial end-to-end instrument, Respiratory Diagcore (Qiagen), or a panel of laboratory-developed real-time PCR assays (organisms tested: SARS-CoV-2, Influenza A + B, Parainfluenza type 1, parainfluenza 1 + 2 + 3, Respiratory Syncytial Virus A/B, Human Metapneumovirus, Adenovirus, Rhinovirus). Respiratory culture was performed using Columbia Blood, Chocolate Blood, MacConkey and Sabouraud agar.

### Sequencing Workflow

The workflow, from host depletion to sequencing, has been described in other papers,^6,8,12^ with reporting following STROBE metagenomics guidance,^13^ with the full protocol available in the online supplement. The following quality controls were used in every run: i) tobacco mosaic virus as an internal control (PC-0107, Leibniz Institute DSMZ-German Collection of Microorganisms and Cell Cultures, Braunschweig, Germany) ii) respiratory swab matrix as a negative control (MC110, Vircell, Granada, Spain) iii) an external positive control containing bacteria, DNA and RNA viruses (NATtrol™ Respiratory Panel 2·1, Zeptometrix, Vernon Hills, IL, USA).

As previously described, prior to nucleic acid extraction, depletion of human DNA was performed using sequential centrifugation at 1200g for 10 minutes, bead-beating with Matrix Lysing D tube (MP biomedicals, Irvine, CA, USA) and endonuclease digestion of released nucleic acid with HL-SAN (ArcticZymes, Tromsø, Norway).

A change to the previously published workflow was made to the extraction method, where 200μl of sample was extracted using the EZ1 Advanced XL with the Virus Mini Kit (Qiagen, Venlo, The Netherlands), excluding carrier RNA, and eluted into 60μl. CDNA was synthesised using LunaScript® RT SuperMix (New England Biolabs, Ipswich, Massachusetts, United States) and double strand synthesis using Sequenase Version 2.0 DNA Polymerase (Applied Biosystems, Waltham, Massachusetts, United States). DNA library preparation was with SQK-RPB004, followed by sequencing for 24 hours on the GridION Mk1 (Oxford Nanopore Technologies, Oxford, UK).

### Bioinformatic Analysis

The bioinformatic analysis was performed using the NHS Metagenomics Platform (https://github.com/GSTT-CIDR/metagenomics_workflow), a containerised toolkit with graphical interfaces, which utilised the classifier Centrifuge^14^ and a curated database composed of elements from FDA-ARGOS, RefSeq and Genbank. FASTQ data was analysed for AMR genes, using Abricate (https://github.com/tseemann/abricate) to screen reads against the CARD^15^ database for matches to AMR genes with a minimum of 90% coverage. Scagaire (https://github.com/quadram-institute-bioscience/scagaire) was used to identify AMR genes known to be associated with clinically relevant organisms.

The Metagenomics Platform generated PDF reports at 0.5, 1, 2, 16 and 24 hours of sequencing. Taxonomic classification outputs were filtered using thresholds determined empirically through ROC analyses. Firstly, classifications were filtered on the Centrifuge ‘score’ (aggregate score of the number of k-mers from a sequencing read which perfect match an organism’s reference score^14^ of 5000 or greater, with viruses alone filtered at 250 or greater. Similar to previous work,^8^ bacteria were reported ‘above threshold’ in the curated sequencing report if they constituted >1% total microbial reads, fungi if five or more reads were detected, and viruses, a single read. Organism specific reporting criteria were based on those developed previously for the workflow.^8^

An orthogonal analysis tool (Organism Query) based on the BLASTn CLI^16^ was included alongside the primary workflow. Leveraging either the ‘nt’ or ‘RefSeq’ databases, the tool extracts a random 50-read subset from binned taxa reported by the primary workflow, and produces a report, summarising hits ranked by bitscore and alignment identity, yielding a ‘most supported organism’ based on a simple majority.

### Clinical Reporting

All clinically reported organisms underwent BLAST query for confirmation unless the pathogen was already known. To facilitate electronic health record (EHR) reporting a bespoke test request and result code were developed. All sequencing reports were first assessed by a microbiologist before reporting to the intensive care teams via call or email and the electronic healthcare record.

Results were reported for clinical staff after 0.5 hours of sequencing (~7 hours after sample receipt) at around 4pm same day and after 16 hours sequencing (24 hours after sample receipt) at 9.00 am the next morning. These time points, different to those used previously for this protocol in the original centre, were chosen based on sensitivity performance in a validation study. The 0.5-hour sequencing results were reported the same-day as a ‘preliminary report’, with the 16-hour sequencing report reported the next morning as a ‘final report’.

The major difference in our reporting criteria from adult settings was to report low level detection of known respiratory pathogens (e.g. Streptococcus pneumoniae) from upper airway samples even if not predominant. This change matched the practice at Great Ormond Street microbiology to report scanty growth of known respiratory pathogens on upper respiratory tract sample cultures. All viruses detected were reported to clinical team. This included herpes viruses, as although not strict respiratory pathogens, the significance of their presence in a paediatric ICU patient could be interpreted and provide utility in the broader clinical context.

Polymerase chain reaction (PCR) was used retrospectively to confirm or refute the presence of pathogens detected by metagenomics but not by routine testing. After analysing the first 115 samples, the threshold for reporting detection of bacteria was changed from >1% of total microbial reads to either >1% of reads or >10 reads. This change was prompted by a re-analysis of the validation study, which showed that it improved sensitivity (0.89 from 0.81) without changing specificity.

As in the previous pilot^8^ a restricted number of acquired antimicrobial resistance genes, including extended-spectrum β lactamases in Enterobacterales, SCCmec in Staphylococcus aureus, and vancomycin-resistance gene clusters in Enterococcus faecium, were analysed and included in the clinical report. An absence of these resistance genes in relevant organisms in the bioinformatics report was not used as evidence of susceptibility, as this is beyond the scope of the bioinformatics pipeline.

### Performance and clinical impact analysis

Results were compared with samples sent for routine respiratory culture and PCR within 24 hours of metagenomics sampling. Where there was discrepancy between metagenomics and culture or PCR, results were confirmed using targeted PCR or 16S rRNA gene sequencing. As with previous publications, we found that orthogonal PCR was highly likely to confirm positive results^6,8^ so if PCR could not be performed (for example if no PCR test for an organism was available), these results were excluded from performance analysis (n=7).

### Clinical impact

Routine data on patient demographics, antimicrobial prescribing, patient mortality, and hospital stay were collected through automated data extraction from the electronic healthcare record. Data on clinical impact of metagenomics were collected retrospectively from the patient electronic healthcare record. Assessment of clinical impact looked at a) impact on antimicrobial prescribing (if there was either a direct referral to mNGS in the patient notes, or an antimicrobial decision was made shortly after a metagenomic result which was congruent with the metagenomic result which differed from culture or PCR) b) pathogen identification not found on routine testing, c) identification of organisms with infection control/public health implications not found on routine testing. All patient notes were reviewed by both a microbiologist and intensive care physician, and any non-concordance being reviewed by a second intensive care physician.

### End-user feedback

To further assess and iteratively improve the implementation of respiratory metagenomics, an anonymous semi-structured questionnaire was carried out with end-users (intensive care staff) (in Supplementary data). This received 33 responses, all ICU staff, including 16 nurses, nine resident doctors, five consultants, and two other staff members.

### Ethics approval

This study evaluates the clinical impact of a newly implemented diagnostic service, with retrospective comparison of results with other pre-existing standard of care diagnostic assays such as PCR and culture. Specific patient consent was not required as metagenomic testing was performed as part of standard of care.

## Results

### Sample composition and demographics

174 samples from 122 patients were tested by metagenomics during a five-month period, from 1^st^ November 2024 to 28th February 2025 on the two ICUs. The samples consisted of 120 endotracheal (ET) secretions, 7 broncho-alveolar lavage samples, 46 nasopharyngeal aspirates (NPAs) and one pleural fluid. Patient demographics and clinical details are shown in Table 1.

**Table 1.** patient demographics and clinical details.

| Metric | Value |
| --- | --- |
| **Patients** | **122** |
| Age (median, IQR) | 0.3 (0.1 - 3.0) |
| Male | 66 (54.1%) |
| Female | 55 (45.9%) |
| Cardiac Intensive Care (CICU) | 60 (49.2%) |
| Paediatric Intensive Care (PICU) | 62 (50.8%) |
| **Samples** | **174** |
| Ventilated | 147 (84.5%) |
| Not Ventilated | 27 (15.5%) |
| On Broad Spectrum Antibiotics | 132 (75.9%) |
| Samples ≥48h after admission | 123 (70.7%) |

In three samples the internal control failed (as evidenced by the absence of tobacco mosaic virus reads) and were subsequently excluded.

### Organism detection

Performance characteristics by kingdom and sequencing duration are shown in Table 2, by kingdom and sample type in Table 3, and by organism group in Figure 1.

**Figure 1.**
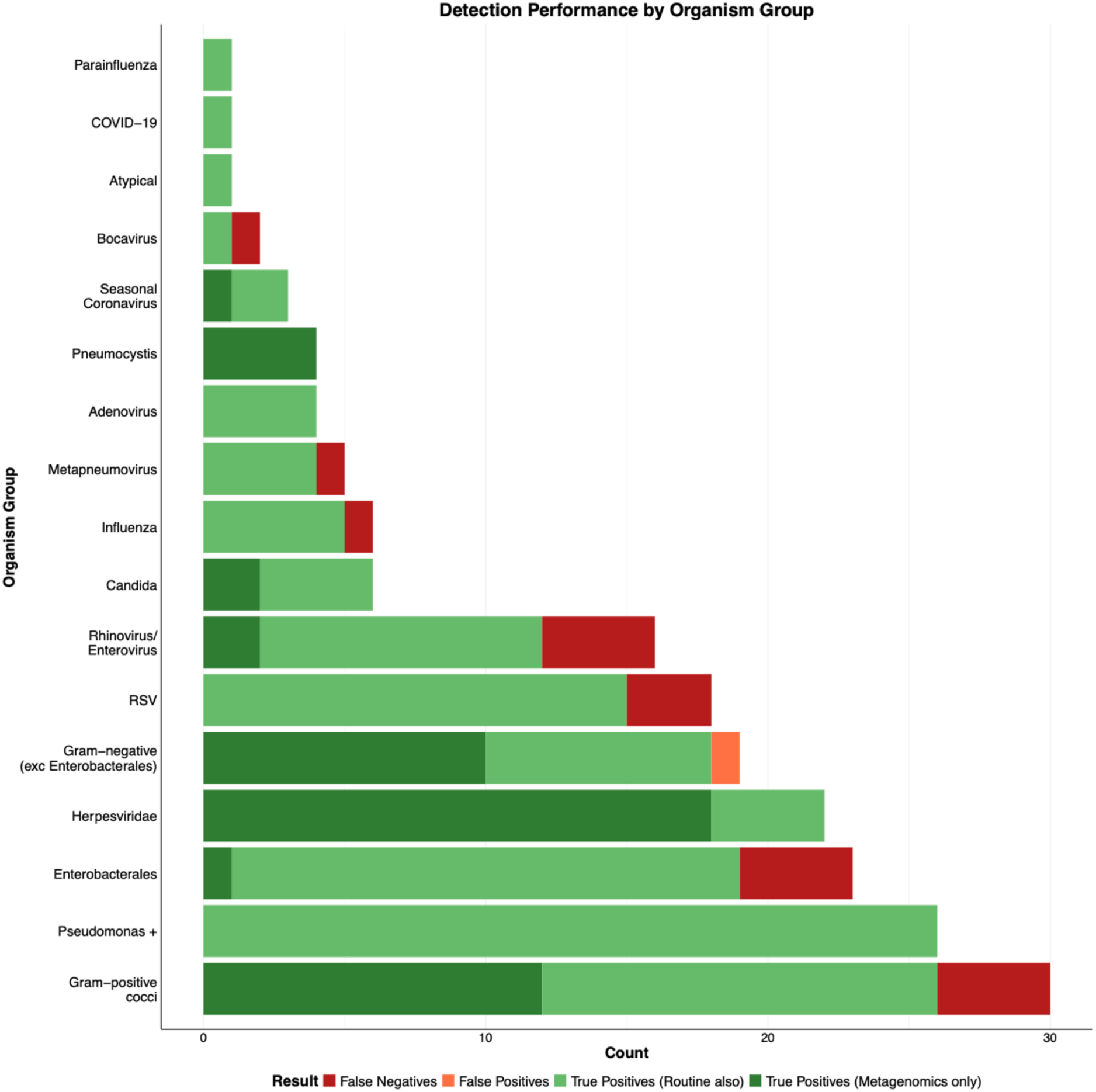
Metagenomics performance characteristics by organism group

**Table 2.** Performance characteristics by kingdom & sequencing duration.

| Kingdom | Timepoint | n | TP | TN | FP | FN | Sensitivity (%) | Specificity (%) | NPV (%) | PPV (%) |
| --- | --- | --- | --- | --- | --- | --- | --- | --- | --- | --- |
| Bacteria | 16 hour | 174 | 67 | 98 | 1 | 8 | 89 (80 – 95) | 99 (95 – 100) | 92 (86 – 97) | 98 (92 – 100) |
| Bacteria | 0.5 hour | 174 | 53 | 98 | 1 | 22 | 71 (59 – 81) | 99 (95 – 100) | 82 (74 – 88) | 98 (90 – 100) |
| Fungi | 16 hour | 174 | 9 | 165 | 0 | 0 | 100 (66 – 100) | 100 (98 – 100) | 100 (98 – 100) | 100 (66 – 100) |
| Fungi | 0.5 hour | 174 | 5 | 165 | 0 | 4 | 56 (21 – 86) | 100 (98 – 100) | 98 (94 – 99) | 100 (48 – 100) |
| Virus | 16 hour | 174 | 58 | 107 | 0 | 9 | 87 (76 – 94) | 100 (97 – 100) | 92 (86 – 96) | 100 (94 – 100) |
| Virus | 0.5 hour | 174 | 37 | 107 | 0 | 30 | 55 (43 – 67) | 100 (97 – 100) | 78 (70 – 85) | 100 (91 – 100) |

**Table 3.**
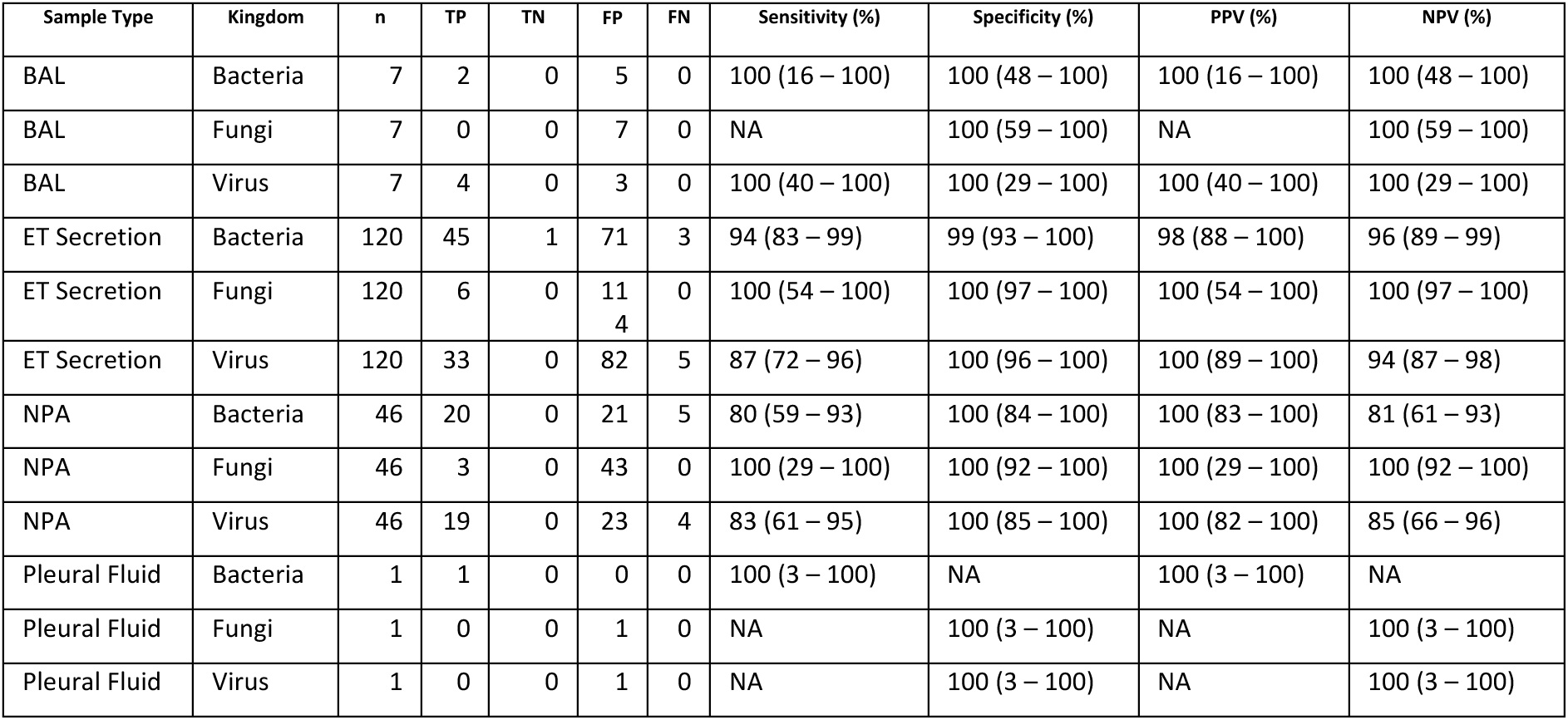
Performance characteristics by kingdom & sample type.

Discordant results are shown in supplementary table 1. There was one false positive compared to routine testing - Haemophilus influenzae in a polymicrobial sample, which was not isolated on culture, and for which targeted PCR was negative. Bacterial false negatives were (1) Scanty growth of Streptococcus pneumoniae in context of heavy growth of Moraxella catarrhalis (which was detected by metagenomics), (2-4) three samples with heavy growth of Staphylococcus aureus cultured in a polymicrobial samples, which were detected by metagenomics but did not meet reporting criteria of >1% of total microbial reads, (6-8) four samples with gram negative bacilli which were scanty growth in culture (two Enterobacter hormaechei and two Klebsiella oxytoca).

There were nine viral false negatives, including Respiratory syncytial virus (n=3, ct values of targeted PCR = 27, 32, 33), Rhinovirus/Enterovirus (n=4, ct = 30, 34, 35, 36), Human metapneumovirus (n=1, ct = 33), Influenza B (n=1, ct = 25) and Bocavirus (n=1, ct = 32.5).

26/174 of samples reported 31 bacteria where prediction of antimicrobial resistance based on target genes could be attempted and there was a culture isolate to compare to (18 Enterobacteriaceae, 11 *Staphylococcus aureus*, and 2 Enterococci). Of these, one had ESBL related genes, and one VRE related genes present, both of which were later confirmed phenotypically in culture. One sample, a *Staphylococcus aureus*, with 52 reads in the final report, did not report resistance (MRSA) which was found in culture.

42/174 samples (24%) returned 50 (26 bacteria, 20 viruses and six fungi) organisms not detected in the routine workflow, which were subsequently confirmed by orthogonal PCR (supplementary table 1).

24/174 samples (14%) had organisms which were not clearly associated with paediatric respiratory disease (Herpesviridae and Pneumocystis jirovecii) in this patient cohort, but the decision was made to clinically report due to potential significance outside the respiratory compartment.

### Clinical Impact

In total, metagenomic findings in 29.3% of samples (51/174) were judged to have had impact on patient management, displayed in figure 2 and table 4. Results influenced antimicrobial prescribing in 26.4% (46/174). 9.2% (16/190) were cessation of antibiotics, 5.7% (10/190) starting antibiotics, 3.4% (6/190) antibiotic change and 8% (14/190) confidence to continue an empirical antibiotic choice. 2.3% of samples (4/190) had potential new infection control or public health implications (three detections of rhinovirus, and one of *Neisseria meningitidis)*. One sample had an impact on diagnosis only (CMV detected leading to detection of a CMV viraemia).

**Figure 2.**
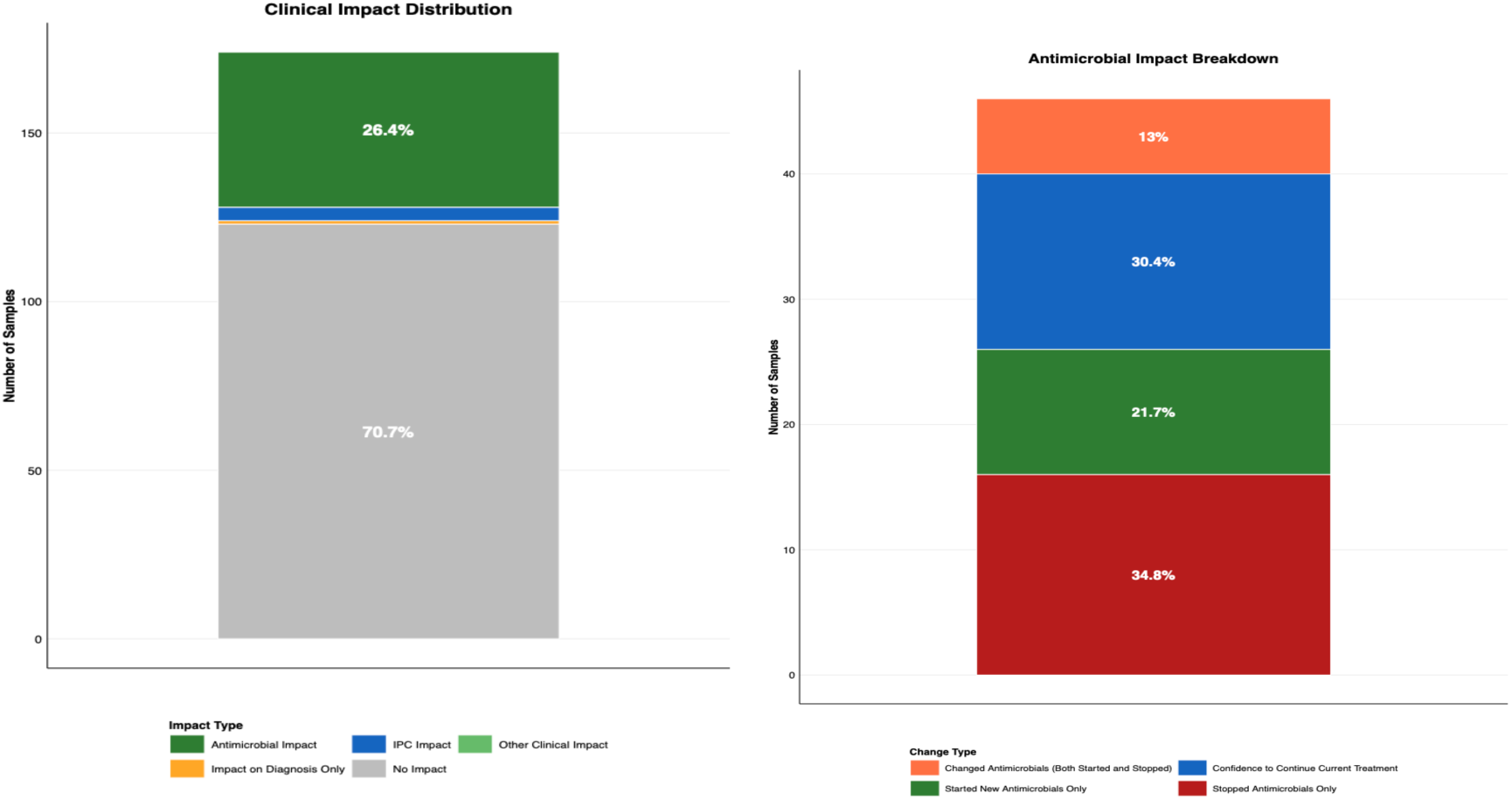
clinical impact of metagenomics and antimicrobial impact breakdown

**Table 4.** clinical impact of metagenomics.

| Measure | Count | Total | Percentage |
| --- | --- | --- | --- |
| Impact on Care (Yes) | 51 | 174 | 29.3% |
| - Impact on Diagnosis Only | 1 | 174 | 0.6% |
| Impact on Antimicrobials (Yes) | 46 | 174 | 26.4% |
| - Started New Antimicrobials Only | 10 | 174 | 5.7% |
| - Stopped Antimicrobials Only | 16 | 174 | 9.2% |
| - Changed Antimicrobials (Both Started and Stopped) | 6 | 174 | 3.4% |
| - Confidence to Continue Current Treatment | 14 | 174 | 8% |
| IPC Intervention Required (Yes) | 4 | 174 | 2.3% |

None of the false negatives, or the false positive, had a direct clinical impact. The culture results for bacteria where metagenomics returned a false negative did not impact patient care directly. None of the viral false negatives had a negative clinical impact, as PCR returned results before metagenomics. In 5/10 (3 RSV, 1 Influenza B, 1 Rhinovirus) of these samples, the PCR had a direct clinical impact which would have been missed if metagenomics alone was used, of which all had IPC implications, and 3/5 (2 RSV, 1 Influenza B) informed clinical diagnosis and management.

### Staff questionnaire results

72% (23/32) of staff thought metagenomics was a positive addition to ICU microbiology testing, 9% (3/32) thought neutral, and 19% (6/32) were unsure, with no staff thinking metagenomics a negative addition, displayed in figure 3. 67% (22/33) thought metagenomics provided additional benefit over current routine testing, with 9% (3/33) seeing no benefit, and 24% (8/33) unsure.

**Figure 3.**
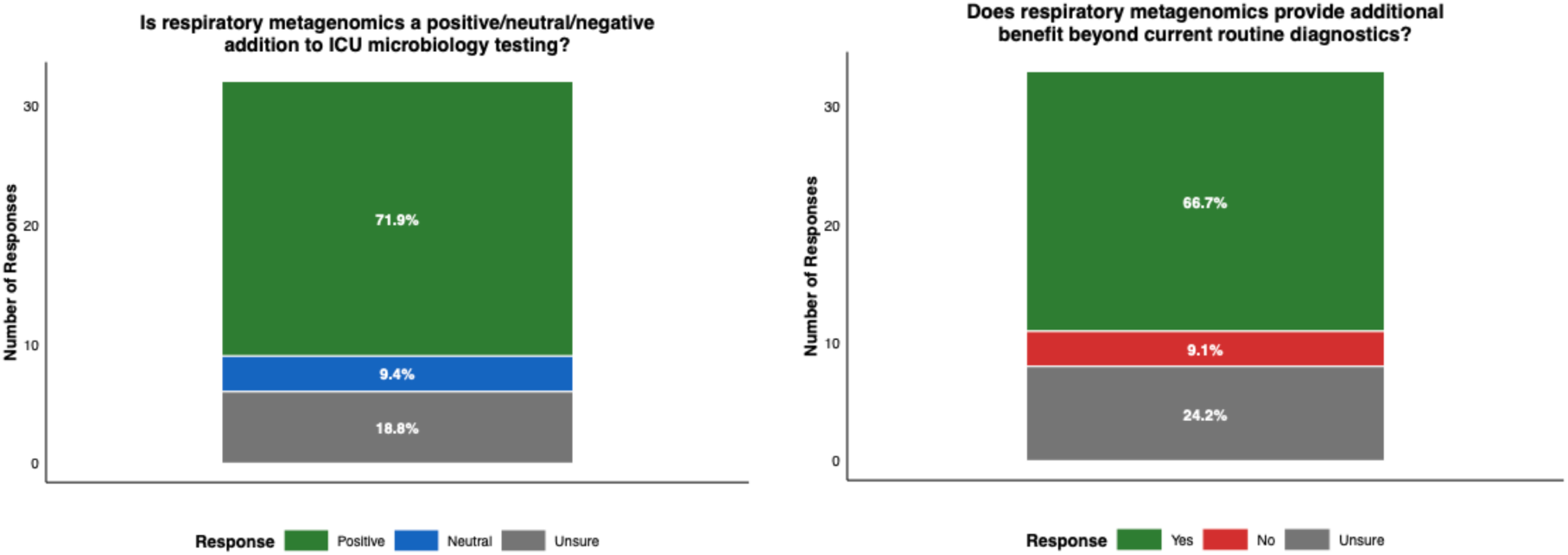
ICU staff views on respiratory metagenomics on anonymous questionnaire

Qualitative questions were asked around likes and dislikes regarding the new service, and suggestions for improvement. Positive comments centered around the rapidity of results and the influence on antimicrobial prescribing, especially in giving confidence to stop antibiotics. Negative comments discussed the difficulty of getting samples to the lab before 9am for same-day results, and on interpreting this novel test, especially in the context of perceived high sensitivity.

## Discussion

We demonstrate the successful implementation of a rapid respiratory metagenomics diagnostic service; utilising protocols transferred outside of the centre where they were developed and applied for the first time to a paediatric intensive care population. Key adaptations compared to the previously described service in adult population have been a shift in sample types with proportionally fewer broncho-alveolar lavages and more endo-tracheal secretions, reflecting local sampling strategy in children, and adaptation of the thresholds for reporting bacterial detections in the context of polymicrobial samples. We also integrated reporting into our electronic healthcare record and evaluated perceptions of the service from clinical ICU staff.

Performance is comparable to previously published work on the protocol,^8^ and the broader field.^17^ Organism detection and performance is comparable to previous metagenomic studies in paediatric intensive care population.^9,11,18,19^ Unlike these studies however, which by and large did not deliver results within a clinically relevant timeframe (and none providing same-day), our study focussed on a rapid service with sample selection by intensivists and minimal exclusion criteria, leading to impact beyond just diagnosis of unexpected organisms. This had led to potential clinical impact (~30%) comparable to previous reports for this protocol, with the largest impact being on antimicrobial prescribing, suggesting a possible role for metagenomics in anti-microbial stewardship.

Very high specificity (>95%) across all three organism kingdoms allowed an excellent positive predictive value, combining with the rapid reporting time to influence intensivists’ prescribing decisions, with impact arising from metagenomic reporting bacteria earlier than culture. Sensitivity for bacteria and fungi was also good, with occasional false negatives arising in polymicrobial or scanty growth samples as previous work^8^. A significant percentage (23%) of metagenomics samples reported results not discovered on routine testing (but confirmed later via PCR), increasing diagnostic yield and contributing to clinical impact.

Negative respiratory metagenomics results, together with full clinical and laboratory findings, provided additional evidence that infection was unlikely therefore allowed intensivists enough confidence to act on wholly negative results – for example stopping antibiotics in unwell children with RSV bronchiolitis. It also gave confidence that there was not a rare, unexpected or atypical cause of infection. Nonetheless the reduced sensitivity of this mNGS method compared to targeted PCR means PCR is still required as standard of care.

Other examples of significant clinical benefit have included first detection of an ESBL Klebsiella in the respiratory compartment, later confirmed by culture, and confirming monomicrobial Group A Streptococcus in a pleural fluid sample from an empyema. Integration of the metagenomics service into routine service delivery, including reporting results via LIMS (Laboratory Information Management System / electronic healthcare record) has been integral to the provision of rapid results.

The pan-microbial detection expands knowledge of the paediatric respiratory microbiome, with increased detection of organism such as Human herpes viruses 6 & 7 and Pneumocystis jirovecii compared to routine testing, findings previously reported in paediatric metagenomic studies.^11,18,19^ Of uncertain significance currently, we hope as this test becomes more established, larger datasets may enable further understanding.

Issues faced in our implementation included the inability to predict antimicrobial sensitivities in most cases beyond a selection of resistance genes, making it harder consistently to influence prescribing in an intensive care population. In this cohort, we also did not discover any truly ‘unexpected’ respiratory pathogens, one of the selling points of metagenomics, and it may be this aspect of metagenomics manifests more in acute hospitals with an A&E department, other body compartments, or is of greater value in a winter season with unexpected emerging pathogens. In addition, despite our assessment suggesting significant clinical impact, it must be acknowledged that this was retrospectively assessed, and the impact of these changes cannot be fully assessed in the absence of a control group.

The high sensitivity for bacteria of the protocol has led to some nuance in communicating the presence of organisms such as Streptococcus pneumoniae Neisseria meningitidis and Haemophilus influenzae in upper respiratory tract samples, an issue previously identified for metagenomic implementation.^5^ Although these difficulties were mitigated by the protocol being embedded with clinical microbiologists, we hope that developing knowledge of the respiratory microbiome, host response^20^ and influence of viral infection^24,20212221^ may allow more precision in future reporting.

This protocol also had difficulty detecting RNA viruses at higher ct values (7/9 of the false negative virus results were ct > 30), findings consistent with other nanopore based protocols^22^, but still important in a paediatric intensive care, as well as for the use of metagenomics as a surveillance tool for emerging pathogens of pandemic potential. The potential impact of metagenomics was reduced by our centre’s use of a rapid multiplex PCR panel (Respiratory Diagcore) as standard of care - the impact may be greater in other centres without rapid (<4 hours TAT) multiplex PCR capability. To replace PCR as gold standard, viral sensitivity will need to improve. There is ongoing work to tackle these issues, both within the protocol,^23^ and via additions such as targeted enrichment panels.^24^

Routinely collected ICU admission and outcome data (supplementary table 2) did not show any significant change for the overall ICU cohort (regardless of metagenomic testing) in average duration of ventilation, organ support or ICU stay, or patient survival, compared to two previous winters. Lack of significant impact on these overall cohort statistics is unsurprising for a diagnostic test of a single organ system given the sample size, but these will be important to analyse in any further studies with control arms.

This implementation, providing a large sample size in a paediatric population, is one of the first demonstrations of adapting an existing clinical metagenomics protocol to a new centre with a different patient population. The rapid turnaround time and strong performance have been maintained, despite changes to sample make-up in a paediatric population. Potential benefit has been seen in impacting antimicrobial prescribing, whilst known issues such as reduced sensitivity for RNA viruses have been more prominent in our cohort. Progressing respiratory metagenomics as a clinical service will involve continuing refinement of the protocol, and multi-centre studies^25^ with control arms to assess true clinical impact and cost-effectiveness.

## Supporting information

ICU Admissions Data

Staff Questionnaire

Full Winter Data-Set

## Data Availability

All data produced in the present study are available upon reasonable request to the authors

