## Supplementary material for "Implementing rapid pan-microbial metagenomics in paediatric intensive care": ICU Admissions Data

| **PICU** | 2022-23 | 2023-24 | 2024-25 |
| --- | --- | --- | --- |
| Number of admissions   - Planned (%) - Unplanned (%) | 391  100 (25.6)  291 (74.4) | 305  95 (31.1)  210 (68.9) | 287  95 (33.1)  192 (66.9) |
| Predicted Index of Mortality %, median (IQR) | 1.6 (0.6-5.3) | 1.6 (0.6-4.3) | 1.0 (0.5-4.0) |
| Admission diagnosis category, n (%)   - Respiratory - Cardiovascular - Neurology - Haem/oncology - Other | 126 (32.2)  17 (4.3)  78 (19.9)  30 (7.7)  140 (35.8) | 95 (31.1)  21 (6.9)  37 (12.1)  17 (5.6)  135 (44.3) | 108 (37.6)  17 (5.9)  19 (6.6)  13 (4.5)  130 (45.2) |
| Length of invasive ventilation in days, median (IQR) | 2 (0-5) | 2 (0-5) | 1 (0-2) |
| Length or organ support in days, median (IQR) | 3 (1-6) | 3 (2-6) | 3 (2-5) |
| Length of ICU stay in days, median (IQR) | 3 (1-6) | 3 (2-6) | 2 (1-5) |
| Survival, n (%) | 370 (94.6) | 280 (91.8) | 262 (93.6)* |

| **CICU** | 2022-23 | 2023-24 | 2024-25 |
| --- | --- | --- | --- |
| Number of admissions   - Planned (%) - Unplanned (%) | 222  143 (64.4)  79 (35.6) | 269  179 (66.5)  90 (33.5) | 213  130 (61.0)  83 (39.0) |
| Predicted Index of Mortality, median (IQR) | 1.3 (0.8-3.9) | 1.2 (0.7-3.7) | 1.2 (0.7-4.5) |
| Admission diagnosis category, n (%)   - Respiratory - Cardiovascular - Other/Unknown | 18 (8.1)  194 (87.4)  10 (4.5) | 32 (11.9)  220 (81.8)  17 (6.3) | 12 (5.6)  132 (62.0)  69 (32.4) |
| Length of invasive ventilation in days, median (IQR) | 2 (1-6) | 2 (1-4) | 2 (1-5) |
| Length or organ support in days, median (IQR) | 2 (4-9) | 3 (2-7) | 4 (3-9) |
| Length of ICU stay in days, median (IQR) | 2 (4-9) | 3 (2-7) | 4 (2-9) |
| Survival, n (%) | 216 (97.3) | 262 (97.4) | 193 (96.0)** |

**Table:** Summary data of general PICU and CICU admissions November-February with the respiratory metagenomic service available (2024-25) and the two years prior (2022-23 and 2023-24).

*outcome data not available from 7 cases – denominator of 280 used for outcome variables.

**outcome data not available for 12 cases denominator of 201 used for outcome variables
