## Supplementary material for "Implementing rapid pan-microbial metagenomics in paediatric intensive care": Staff Questionnaire

### Respiratory Metagenomics Staff Questionnaire

1. 1. a) What is your staff role

   *Mark only one oval.*

   - Consultant
   - Resident Doctor
   - Nurse
   - Allied Healthcare Professional
   - Other

2. 1. b) What is your specialty?

   *Mark only one oval.*

   - Microbiology/Virology
   - Infectious Diseases
   - Intensive Care
   - Other

3. 2. a) Have you been involved in **deciding to send** a respiratory metagenomics sample?

   *Mark only one oval.*

   - Yes
   - No

4. 2. b) **If yes to sending**, are you clear when it is appropriate to send?

*Mark only one oval.*

- Yes
- No

5. 2. c) **If yes to sending**, any suggestions for change in policy around sending metagenomic?

_______________________________

6. 3. a) Have you been involved in **taking** a respiratory metagenomics sample?

*Mark only one oval.*

- Yes
- No

7. 3. b) **If yes to taking**, did you have any issues taking and sending the sample?

*Mark only one oval.*

- Yes
- No

8. 3. c) **If yes to taking**, please detail any issues with taking, and any suggestions for change?

_______________________________
_______________________________
_______________________________
_______________________________
_______________________________

9. 4. a) Have you been involved in **actioning the results** of a respiratory metagenomics sample?

*Mark only one oval.*

- Yes
- No

10. 4. b) **If yes to actioning,** did you find the EPIC report easy to understand?

*Mark only one oval.*

- Yes
- No

11. 4. c) **If yes to actioning,** how do you find out results?

*Mark only one oval.*

- EPIC review
- Daily emails
- Discussion in AMS rounds
- Other

12. 4. c) **If yes to actioning,** did it impact your patient management?
Please describe how if so (ie. broadening/narrowing/stopping antibiotics, confirming antibiotic choice, infection control, helping clinical diagnosis)

13. 5. a) Do you think respiratory metagenomics is a positive, neutral or negative addition to ICU microbiology testing (or unsure)?

*Mark only one oval.*

- Positive
- Neutral
- Negative
- Unsure

14. 5. b) Do you think respiratory metagenomics provides additional benefit beyond current routine diagnostics (eg. respiratory diagcore pcr and respiratory culture)

*Mark only one oval.*

- Yes
- No
- Unsure

15. 6. Please describe anything you particularly like or dislike about respiratory metagenomics (optional)

16. 7. Please suggest any changes to the respiratory metagenomics pathway (optional)

This content is neither created nor endorsed by Google.

Google Forms
